# Postoperative glycemic variability as a predictor for one-year mortality following coronary artery bypass grafting: A retrospective cohort study

**DOI:** 10.1101/2025.02.13.25322258

**Authors:** Jiasen Ma, Junjun Qin, Fengkai Mao, Changlin He, Daqing Ma, Jinpiao Zhu

## Abstract

**Background:** Increased postoperative glycemic variability (GV) has been shown to be associated with an increased risk for adverse outcomes. However, the impact of postoperative GV on the long-term mortality after cardiac surgery, particularly with regarding other associated interactive factors, remains unknown. In this study, we sought to investigate whether postoperative GV is associated with one-year mortality, and to determine whether preoperative hemoglobin A1C (HbA1C) levels or diabetes status contribute to the risk of GV-related one-year mortality following coronary artery bypass grafting (CABG).

**Methods:** Data from the Medical Information Mart for Intensive Care-IV (MIMIC-IV) database derived from 3,598 patients, who underwent their CABG surgery and subsequently admitted to the ICU between 2008 and 2019, were retrospectively analysed. Patients were categorized into two groups based on the postoperative GV levels: the GV-Low group (GV < 0.219, n=2,399) and the GV-High group (GV ≥ 0.219, n=1,199). Cox proportional hazards models were used to examine the association between postoperative GV and one-year mortality. Kaplan-Meier curves assessed cumulative survival, while restricted cubic splines (RCS) evaluated potential non-linear relationships. Cox regression was carried out to determine the impact of postoperative GV changes on one-year mortality.

**Results:** The one-year mortality rate was 4.92% following CABG. Multivariate Cox proportional hazards analysis revealed a significant association between postoperative GV and one-year mortality, with harzard ratio of 10.372 (95% CI, 3.973–27.077) in the unadjusted model, 9.065 (95% CI, 3.387–24.263) in the partially adjusted model, and 4.599 (95% CI, 1.662–12.726) in the fully adjusted model. This association followed a linear pattern (*p*-non-linearity = 0.962). In addition, the association was stronger in patients without diabetes millutes (DM) (odds ratio [OR], 2.781; 95% CI, 1.709–4.526) or in patients with preoperative HbA1c levels ≤ 7.0% (OR, 2.225; 95% CI, 1.537–3.221).

**Conclusion:** Increased postoperative GV is associated with higher one-year mortality following CABG, particularly in patients without DM. These findings may offer valuable guidance for targeted strategies to reduce mortality after cardiac surgery.

## Introduction

Coronary artery bypass grafting (CABG) is an effective treatment for improving life quality and prolonging lifetime in patients with coronary artery disease (CAD), particularly those with complex multivessel disease ^1,2^. While advancements in surgical techniques and pharmacological treatments improve survival, the challenge of mitigating postoperative complications, including mortality, persists ^3,4^. Therefore, it is essential to identify reliable and practical mortality predictors to optimize patient management and improve prognosis following CABG.

Glycemic variability (GV) is a key indicator of glycemic fluctuations, reflecting the degree of metabolic instability in critically Ill patients ^5^. Previous studies showed that increasing GV conferred as an independent risk of mortality in a heterogeneous patient population ^6,7^. Unlike persistent hyperglycemia, GV represents acute and dynamic blood glucose fluctuations, often driven by postoperative stress-induced hyperglycemia and systemic inflammatory responses ^8^. Early study indicated that increased GV in patients with elevated preoperative hemoglobin A1C (HbA1C) may predict major adverse outcomes (MAEs) within 30 days following CABG ^9^. In contrast, another study demonstrated no significant association between preoperative HbA1c or postoperative GV and MAEs in patients undergoing isolated cardiac valvular surgery during the 30-day postoperative period ^10^. Therefore, the larger-scale population studies are needed to further validate whether the postoperative GV can be treated as a predictor for the MAEs, particularly the long-term mortality, following cardiac surgery.

In the current study, we investigated whether postoperative GV is associated with the risk of postoperative one-year mortality after CABG surgery, and whether preoperative HbA1C levels or diabetes status can enhance this risk.

## Methods

### Data source

The data were obtained from the Medical Information Mart for Intensive Care-IV (MIMIC-IV) database (version 3.0), comprising more than 65,000 patients admitted to the ICU between 2008 and 2019 ^11^. As this study only analyzed de-identified data, the requirement for informed consent was waived. The first author (J.M.) completed an online course provided by the National Institutes of Health and passed the Human Research Participant Protection Examination, obtaining permission for the Collaborative Institutional Training Initiative and access to the MIMIC-IV database (certification number: 64145697).

### Study population

Patients were enrolled if they met the following criteria: (1) Undergoing the first CABG surgery; (2) The first-time admission to the ICU; (3) With ≥ 3 blood glucose measurements in the first 72-h postoperative period; (4) Age ≥ 18 years old. Ultimately, a total of 3,598 patients were included in this study (Figure 1).

**Figure 1.**
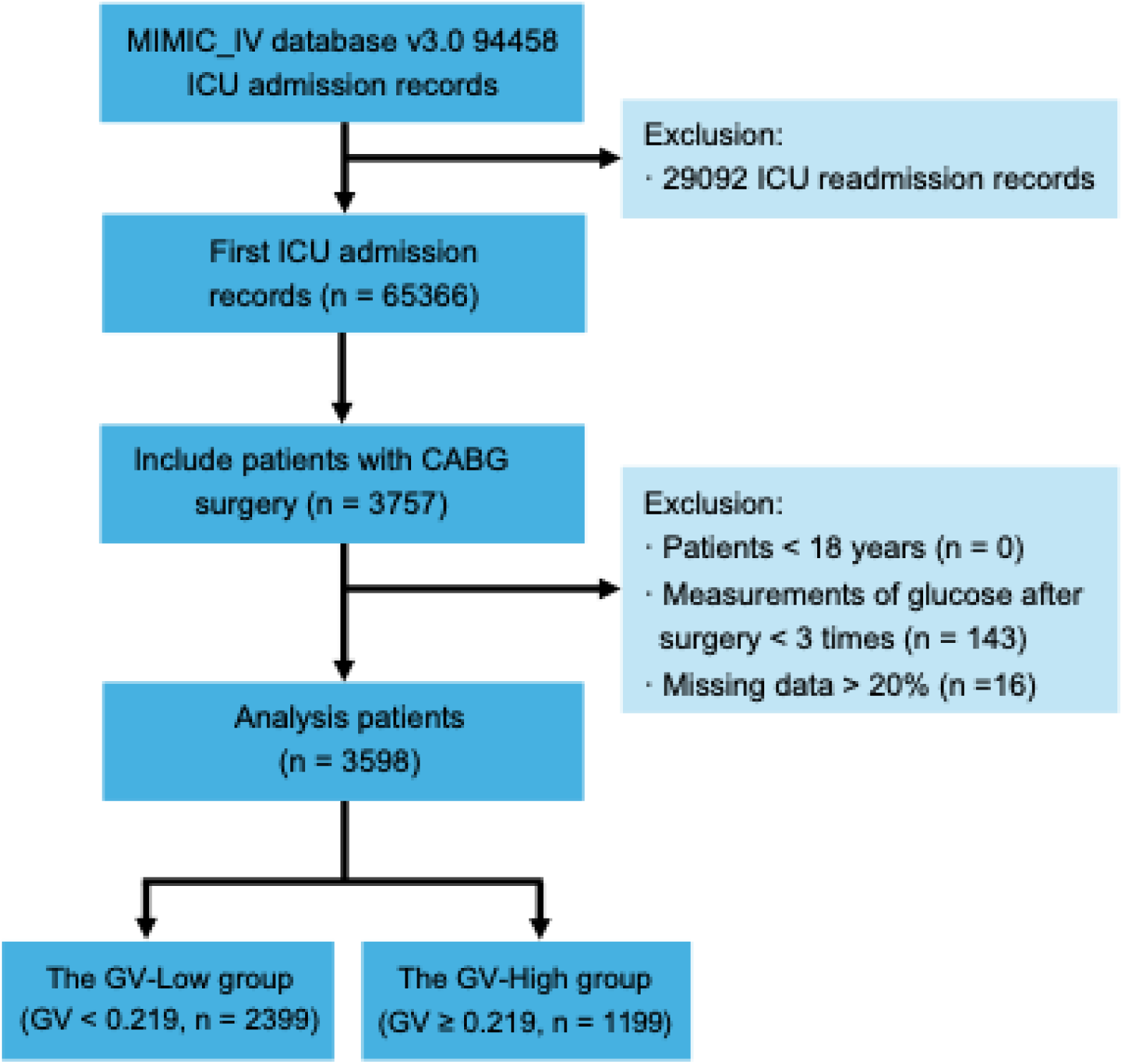
Flow diagram of patient selection. ICU, intensive care unit; CABG, coronary artery bypass grafting; GV glycemic variability.

### Data extraction and definitions

The data of demographic characteristics, vital signs, comorbidities, laboratory measurements, and treatments during hospitalization were extracted and performed using the Navicat Premium software (version 17.0.14, PremiumSoft CyberTech Ltd.) with a structured query language (SQL). Gemographic information included age, gender, body mass index (BMI) and race background. Vital signs comprised the 24-h postoperative averages for heart rate, systolic blood pressure (SBP), diastolic blood pressure (DBP) and peripheral capillary oxygen saturation (SpO₂). Comorbidities included hypertension, diabetes mellitus (DM), acute myocardial infarction (AMI), previous MI, heart failure, chronic obstructive pulmonary disease (COPD), chronic kidney disease (CKD), cerebrovascular disease and the Charlson Comorbidity Index (CCI). Laboratory measurments were hemoglobin, white blood cell (WBC) count, platelet count, glycosylated hemoglobin (HbA1c), serum creatinine (Scr), blood urea nitrogen (BUN), albumin, serum potassium and serum sodium before and after CABG. Also, plasma glucose measurements were done three times within the initial 72-h post-CABG. The estimated glomerular filtration rate (eGFR) was calculated using the Modification of Diet of Renal Disease equation^12^. Postoperative treatments included renal replacement therapy (RRT), duration of mechanical ventilation, and medication therapy. Additionally, the number of coronary artery bypass grafts and the Sequential Organ Failure Assessment (SOFA) scores were recorded at 24-h postoperative period. Patients with over 20% missing data were excluded for analysis. For variables with less than 20% missing, multiple imputation was performed using the random forest method provided by the mice package in R software (version 4.4.1).

### Study grouping and outcome endpoint

Glycemic variability (GV) was presented as the coefficient of variation, defined as the ratio of the standard deviation to the mean of the first three plasma glucose measurements during postoperative period ^7,13^. The 3,598 patients were devided into two groups based on their GV values in a 2:1 ratio: the GV-Low group (GV < 0.219) and the GV-High group (GV ≥ 0.219). The primary outcome was one-year postoperative survival status.

### Statistical analysis

Continuous variables were presented as medians with interquartile ranges (IQR) or mean ± standard deviation (SD), while categorical variables were presented as frequencies and percentages. Group differences were assessed using the Kruskal-Wallis rank sum test for continuous variables and the Chi-squared test for categorical variables. To correct multiple test’s false positive, the Benjamini-Hochberg procedure was applied to control the false discovery rate (FDR).

Kaplan-Meier curves were used to calculate the cumulative incidence of one-year postoperstive mortality for both groups, and the comparisons between both groups were done with the log-rank test. Cox proportional regression models were employed to estimate the hazard ratios (HR) and 95% confidence interval (CI) of GV for one-year mortality following CABG. Stepwise regression was applied to adjust for variables with *p* < 0.10 invariable regression analysis, along with clinically relevant variables. Multicollinearity among the selected variables was evaluated using variance inflation factor analysis, ensuring that all included variables had acceptable values (≤ 10).

Restricted cubic spline (RCS) models were conducted to examine the linearity of the relationship between the GV and the postoperative one-year mortality. These models were adjusted for potential confounders to ensure robustness. Subgroup and interaction analyses were performed based on DM status and preoperative HbA1c levels to further explore the independent and interactive effects of GV on the one-year mortality. Forest plots were used to visually present the results of subgroup and interaction analyses.

Model performance and complexity were evaluated using the Akaike Information Criterion (AIC) and Bayesian Information Criterion (BIC), with lower values indicating better model fit and balance. Sensitivity analysis was carried out with a median-based approach, where GV was dichotomized into high and low groups based on the median value to validate the stability of the findings. A statistical significance was defined as a *p*-value less than 0.05.

## Results

### Baseline characteristics of patients undergoing CABG

A total of 3,598 patients who underwent the coronary artery bypass grafting (CABG) was enrolled in the study. The patients were divided into two groups based on their postoperative glucose variability (GV): the GV-Low group (GV < 0.219, n=2,399) and the GV-High group (GV ≥ 0.219, n=1,199) (Figure 1). Baseline characteristics of patients before CABG were detailed in Table 1. The age of the participants was over 69.08 ± 10.01 years old. The overall proportion of male patients was high across the study population, but it was significantly higher in the GV-Low group compared to the GV-High group (82.28% vs 75.06%, *p* = 0.001). Patients in the GV-High group had a higher incidence of acute myocardial infarction (AMI), heart failure, hypertension, diabetes mellitus (DM), chronic kidney disease (CKD) and cerebrovascular disease, and had higher Charlson Comorbidity Index (CCI). In addition, they exhibited increased levels of the glycosylated hemoglobin (HbA1c), serum creatinine (Scr) and blood urea nitrogen (BUN), alongside decreased levels of the hemoglobin, albumin and estimated glomerular filtration rate (eGFR). The GV-High group also had higher rates of receiving therapies such as anti-hypertensives including alpha-blockers, antidiabetic medications including insulin and other agents, anticoagulants, and renal replacement therapy (RRT). At a one-year follow-up, the mortality rate in the GV-High group was significantly higher at 6.84%, compared to 3.96% in the GV-Low group.

**Table 1.** Baseline characteristics of patients.

| Variables | Overall (n=3598) | GV-Low (n=2399) | GV-High (n=1199) | p-value | q-value<sup>a</sup> |
| --- | --- | --- | --- | --- | --- |
| **Demographics** |  |  |  |  |  |
| Age, years | 69.08 (61.88, 75.61) | 69.03 (61.79, 75.40) | 69.44 (62.10, 76.22) | 0.280 | 1 |
| Male, n (%) | 2874 (79.88) | 1974 (82.28) | 900 (75.06) | 0.001 | < 0.001 |
| BMI, kg/m<sup>2</sup> | 28.55 (25.34, 32.41) | 28.52 (25.43, 32.25) | 28.60 (25.15, 32.57) | 0.986 | 1 |
| RACE, n (%) |  |  |  | 0.045 | 0.135 |
| White | 2415 (67.12) | 1647 (68.65) | 768 (64.05) |  |  |
| Black | 154 (4.28) | 95 (3.96) | 59 (4.92) |  |  |
| Asian | 90 (2.50) | 50 (2.08) | 40 (3.33) |  |  |
| Other | 939 (26.10) | 607 (25.30) | 332 (27.69) |  |  |
| **Vital signs** |  |  |  |  |  |
| SBP, mmHg | 110.59 (106.22, 115.44) | 110.53 (106.00, 115.50) | 110.77 (106.50, 115.31) | 0.645 | 1 |
| DBP, mmHg | 57.50 (52.80, 61.92) | 58.19 (53.67, 62.58) | 56.00 (51.81, 60.17) | < 0.001 | < 0.001 |
| Heart rate, bpm | 79.64 (74.67, 85.33) | 79.36 (74.46, 84.55) | 80.07 (74.95, 86.71) | 0.001 | 0.063 |
| SpO<sub>2</sub>, % | 98.60 (97.54, 99.39) | 98.55 (97.54, 99.37) | 98.67 (97.53, 99.42) | 0.220 | 1 |
| **Comorbidities** |  |  |  |  |  |
| AMI, n (%) | 1190 (33.07) | 740 (30.85) | 450 (37.53) | < 0.001 | 0.007 |
| Previous MI, n (%) | 123 (3.42) | 75 (3.13) | 48 (4.00) | 0.135 | 1 |
| Heart failure, n (%) | 661 (18.37) | 379 (15.80) | 282 (23.52) | < 0.001 | < 0.001 |
| Hypertension, n (%) | 3111 (86.46) | 2042 (85.12) | 1069 (89.16) | 0.001 | < 0.001 |
| DM, n (%) | 1637 (45.50) | 766 (31.93) | 871 (72.64) | < 0.001 | < 0.001 |
| COPD, n (%) | 568 (15.79) | 378 (15.76) | 190 (15.85) | 0.992 | 1 |
| CKD, n (%) | 778 (21.62) | 411 (17.13) | 367 (30.61) | < 0.001 | < 0.001 |
| Cerebrovascular disease, n (%) | 365 (10.14) | 219 (9.13) | 146 (12.18) | 0.017 | 0.934 |
| CCI | 4.00 (3.00, 6.00) | 4.00 (3.00, 6.00) | 5.00 (4.00, 7.00) | < 0.001 | < 0.001 |
| **Laboratory measurements** |  |  |  |  |  |
| HbA<sub>1c</sub>, % | 6.00 (5.50, 7.20) | 5.80 (5.40, 6.40) | 7.00 (6.00, 8.30) | < 0.001 | < 0.001 |
| Hemoglobin, g/dL | 13.10 (11.70, 14.30) | 13.30 (12.00, 14.40) | 12.50 (11.10, 13.90) | < 0.001 | < 0.001 |
| WBC, 10<sup>9</sup>/L | 7.40 (6.20, 9.00) | 7.40 (6.10, 8.90) | 7.60 (6.30, 9.20) | 0.044 | 1 |
| Platelets, 10<sup>9</sup>/L | 205.00 (170.00, 248.00) | 204.00 (169.00, 246.00) | 208.00 (172.00, 252.00) | 0.137 | 1 |
| Scr, mg/dL | 1.00 (0.80, 1.20) | 1.00 (0.80, 1.10) | 1.00 (0.80, 1.30) | < 0.001 | < 0.001 |
| BUN, mg/dL | 18.00 (14.00, 23.00) | 17.00 (14.00, 22.00) | 19.00 (15.00, 25.00) | < 0.001 | < 0.001 |
| eGFR, mL/min/1.73m<sup>2</sup> | 75.62 (57.72, 90.17) | 78.21 (61.22, 90.82) | 70.37 (51.39, 88.28) | < 0.001 | < 0.001 |
| Albumin, g/dL | 4.00 (3.70, 4.30) | 4.10 (3.80, 4.40) | 4.00 (3.60, 4.20) | < 0.001 | < 0.001 |
| Sodium, mmol/L | 139.00 (137.00, 141.00) | 139.00 (137.00, 141.00) | 139.00 (137.00, 141.00) | 0.004 | 0.243 |
| Potassium, mmol/L | 4.20 (4.00, 4.50) | 4.20 (4.00, 4.50) | 4.20 (4.00, 4.50) | 0.062 | 1 |
| SOFA score | 3.00 (1.00, 4.00) | 3.00 (1.00, 4.00) | 3.00 (1.00, 4.00) | 0.531 | 1 |
| **Treatment during hospitalization** |  |  |  |  |  |
| ACEIs/ARBs, n (%) | 318 (8.84) | 218 (9.09) | 110 (9.17) | 0.528 | 1 |
| Alpha-blockers, n (%) | 989 (27.49) | 600 (25.01) | 389 (32.44) | < 0.001 | < 0.001 |
| Beta-blockers, n (%) | 2966 (82.43) | 1954 (81.45) | 1012 (84.40) | 0.080 | 1 |
| Calcium channel blockers, n (%) | 3474 (96.55) | 2314 (96.46) | 1160 (96.75) | 0.546 | 1 |
| Vasoactive drugs, n (%) | 3413 (94.86) | 2279 (95.00) | 1134 (94.58) | 0.744 | 1 |
| Diuretics, n (%) | 3118 (86.66) | 2071 (86.33) | 1047 (87.32) | 0.710 | 1 |
| Anti arrhythmic drugs, n (%) | 715 (19.87) | 465 (19.38) | 250 (20.85) | 0.025 | 1 |
| Insulin, n (%) | 3476 (96.61) | 2298 (95.79) | 1178 (98.25) | < 0.001 | 0.016 |
| Other antidiabetic drugs, n (%) | 200 (5.56) | 69 (2.88) | 131 (10.93) | < 0.001 | < 0.001 |
| Glucocorticoids, n (%) | 263 (7.31) | 181 (7.54) | 82 (6.84) | 0.689 | 1 |
| NSAIDs, n (%) | 3547 (98.58) | 2366 (98.62) | 1181 (98.50) | 0.158 | 1 |
| Antiplatelets, n (%) | 54 (1.50) | 30 (1.25) | 24 (2.00) | 0.206 | 1 |
| Anticoagulants, n (%) | 923 (25.65) | 572 (23.84) | 351 (29.27) | 0.001 | 0.074 |
| Opioids, n (%) | 3449 (95.86) | 2300 (95.87) | 1149 (95.83) | 0.308 | 1 |
| Sedatives, n (%) | 3461 (96.19) | 2313 (96.42) | 1148 (95.75) | 0.601 | 1 |
| Statins, n (%) | 3146 (87.44) | 2104 (87.70) | 1042 (86.91) | 0.514 | 1 |
| H<sub>2</sub>-receptor antagonists, n (%) | 3212 (89.27) | 2144 (89.37) | 1068 (89.07) | 0.595 | 1 |
| Ventilation, days | 12.00 (6.97, 20.00) | 12.00 (7.00, 19.75) | 12.00 (6.75, 20.89) | 0.781 | 1 |
| RRT, n (%) | 166 (4.61) | 82 (3.42) | 84 (7.01) | < 0.001 | < 0.001 |
| Number of grafts | 1.00 (1.00, 2.00) | 1.00 (1.00, 2.00) | 1.00 (1.00, 2.00) | 0.499 | 1 |
| **Outcomes** |  |  |  |  |  |
| one-year mortality, n (%) | 177 (4.92) | 95 (3.96) | 82 (6.84) | < 0.001 | 0.042 |

Data were presented as median [25 percentile/75 percentile] or n (%). <sup>a</sup> Bonferroni correction for multiple testing. CABG, coronary artery bypass grafting; BMI, body mass index; SBP, systolic blood pressure; DBP, diastolic blood pressure; SpO<sub>2</sub>, peripheral capillary oxygen saturation; MI, myocardial infarction; AMI, acute myocardial infarction; DM, diabetes mellitus; COPD, chronic obstructive pulmonary disease; CKD, chronic kidney disease; CCI, Charlson Comorbidity Index; HbA1c, hemoglobin A1c; WBC, white blood cell; Scr, serum creatinine; BUN, blood urea nitrogen; eGFR, estimated glomerular filtration rate; SOFA, Sequential Organ Failure Assessment; ACEI, angiotensin converting enzyme inhibitor; ARB, angiotensin II receptor blocker; RRT, renal replacement therapy.

### Postoperative outcomes of patients following CABG

At post-one year of CABG, 3,421 patients survived, while 177 patients died (Table 2). Non-survivors were older with less males compared to survivors. They were more likely to have comorbidities such as AMI, previous MI, heart failure, hypertension, DM, CKD and cerebrovascular diseases. In addition, non-survivors exhibited also lower preoperative levels of hemoglobin, albumin and sodium, along with high white blood cell (WBC), Scr, BUN, eGFR and potassium. Furthermore, non-survived patients were treated with fewer antihypertensive and sedative drugs but required more use of vasoactive and antiarrhythmic drugs. They were also needed longer durations of mechanical ventilation and more frequent RRT.

**Table 2.** Characteristics of survivors and non-survivors post one-year CABG.

| Variables | Survivor (n=3421) | Non-survivor (n=177) | p-value | q-value<sup>a</sup> |
| --- | --- | --- | --- | --- |
| **Demographics** |  |  |  |  |
| Age, years | 68.90 (61.71, 75.36) | 73.33 (64.67, 80.94) | < 0.001 | < 0.001 |
| Male, n (%) | 2753 (80.47) | 121 (68.36) | < 0.001 | 0.007 |
| BMI, kg/m<sup>2</sup> | 28.58 (25.40, 32.38) | 27.97 (24.48, 32.12) | 0.119 | 1 |
| RACE, n (%) |  |  | 0.126 | 1 |
| White | 2296 (67.11) | 119 (67.23) |  |  |
| Black | 141 (4.12) | 13 (7.34) |  |  |
| Asian | 88 (2.57) | 2 (1.13) |  |  |
| Other | 896 (26.19) | 43 (24.29) |  |  |
| **Vital signs** |  |  |  |  |
| SBP, mmHg | 110.77 (106.33, 115.50) | 109.19 (104.44, 114.09) | 0.005 | 0.298 |
| DBP, mmHg | 57.67 (53.08, 62) | 53.18 (48.64, 57.73) | < 0.001 | < 0.001 |
| Heart rate, bpm | 79.50 (74.59, 85.14) | 80.50 (76.40, 87.72) | 0.001 | 0.067 |
| SpO<sub>2</sub>, % | 98.58 (97.54, 99.39) | 98.69 (97.40, 99.5) | 0.865 | 1 |
| **Comorbidities** |  |  |  |  |
| AMI, n (%) | 1097 (32.07) | 93 (52.54) | < 0.001 | < 0.001 |
| Previous MI, n (%) | 105 (3.07) | 16 (9.04) | < 0.001 | 0.001 |
| Heart failure, n (%) | 583 (17.04) | 78 (44.07) | < 0.001 | < 0.001 |
| Hypertension, n (%) | 2947 (86.14) | 164 (92.66) | 0.019 | 1 |
| DM, n (%) | 1531 (44.75) | 106 (59.89) | < 0.001 | 0.006 |
| COPD, n (%) | 524 (15.32) | 44 (24.86) | 0.001 | 0.057 |
| CKD, n (%) | 690 (20.17) | 88 (49.72) | < 0.001 | < 0.001 |
| Cerebrovascular disease, n (%) | 325 (9.50) | 40 (22.60) | < 0.001 | < 0.001 |
| CCI | 4.00 (3.00, 6.00) | 7.00 (5.00, 9.00) | < 0.001 | < 0.001 |
| **Laboratory measurements** |  |  |  |  |
| GV | 0.16 (0.09, 0.25) | 0.20 (0.12, 0.31) | < 0.001 | 0.003 |
| HbA<sub>1c</sub>, % | 6.00 (5.50, 7.20) | 6.40 (5.58, 7.60) | 0.168 | 1 |
| Hemoglobin, g/dL | 13.20 (11.80, 14.30) | 11.00 (9.60, 12.70) | < 0.001 | < 0.001 |
| WBC, 10<sup>9</sup>/L | 7.40 (6.20, 9.00) | 8.10 (6.90, 10.20) | < 0.001 | 0.001 |
| Platelets, 10<sup>9</sup>/L | 205.00 (170.00, 247.00) | 202.00 (159.00, 252.00) | 0.323 | 1 |
| Scr, mg/dL | 1.00 (0.80, 1.20) | 1.20 (0.90, 1.90) | < 0.001 | < 0.001 |
| BUN, mg/dL | 18.00 (14.00, 23.00) | 23.00 (17.00, 37.00) | < 0.001 | < 0.001 |
| eGFR, mL/min/1.73m<sup>2</sup> | 76.74 (59.62, 90.44) | 50.84 (31.22, 77.55) | < 0.001 | < 0.001 |
| Albumin, g/dL | 4.00 (3.80, 4.40) | 3.80 (3.30, 4.10) | < 0.001 | < 0.001 |
| Sodium, mmol/L | 139.00 (137.00, 141.00) | 138.00 (135.00, 140.00) | < 0.001 | < 0.001 |
| Potassium, mmol/L | 4.20 (4.00, 4.50) | 4.40 (4.00, 4.70) | 0.001 | 0.050 |
| SOFA score | 3.00 (1.00, 4.00) | 3.00 (1.00, 5.00) | 0.016 | 0.901 |
| **Treatment during hospitalization** |  |  |  |  |
| ACEIs/ARBs, n (%) | 301 (8.80) | 17 (9.60) | 0.816 | 1 |
| Alpha-blockers, n (%) | 926 (27.07) | 63 (35.59) | 0.017 | 0.958 |
| Beta-blockers, n (%) | 2837 (82.93) | 129 (72.88) | < 0.001 | 0.051 |
| Calcium channel blockers, n (%) | 3313 (96.84) | 161 (90.96) | < 0.001 | 0.004 |
| Vasoactive drugs, n (%) | 3257 (95.21) | 156 (88.14) | < 0.001 | 0.004 |
| Diuretics, n (%) | 2986 (87.28) | 132 (74.58) | < 0.001 | < 0.001 |
| Anti arrhythmic drugs, n (%) | 632 (18.47) | 83 (46.89) | < 0.001 | < 0.001 |
| Insulin, n (%) | 3312 (96.81) | 164 (92.66) | 0.006 | 0.322 |
| Other antidiabetic drugs, n (%) | 188 (5.50) | 12 (6.78) | 0.576 | 1 |
| Glucocorticoids, n (%) | 263 (7.31) | 24 (13.56) | 0.002 | 0.100 |
| NSAIDs, n (%) | 3378 (98.74) | 169 (95.48) | 0.001 | 0.065 |
| Antiplatelets, n (%) | 51 (1.49) | 3 (1.69) | 0.748 | 1 |
| Anticoagulants, n (%) | 807 (23.59) | 116 (65.54) | < 0.001 | < 0.001 |
| Opioids, n (%) | 3288 (96.11) | 161 (90.96) | 0.002 | 0.089 |
| Sedatives, n (%) | 3300 (96.46) | 161 (90.96) | < 0.001 | 0.024 |
| Statins, n (%) | 2991 (87.43) | 155 (87.57) | 1 | 1 |
| H<sub>2</sub>-receptor antagonists, n (%) | 3059 (89.42) | 153 (86.44) | 0.261 | 1 |
| Ventilation, days | 12.00 (6.78, 19.50) | 19.00 (8.42, 28.00) | < 0.001 | < 0.001 |
| RRT, n (%) | 108 (3.16) | 58 (32.77) | < 0.001 | < 0.001 |
| Number of grafts | 1.00 (1.00, 2.00) | 1.00 (1.00, 2.00) | 0.961 | 1 |

Data were presented as median [25 percentile/75 percentile] or n (%). <sup>a</sup> Bonferroni correction for multiple testing. CABG, coronary artery bypass grafting; BMI, body mass index; SBP, systolic blood pressure; DBP, diastolic blood pressure; SpO<sub>2</sub>, peripheral capillary oxygen saturation; MI myocardial infarction; AMI, acute myocardial infarction; DM, diabetes mellitus; COPD, chronic obstructive pulmonary disease; CKD, chronic kidney disease; CCI, Charlson Comorbidity Index; HbA1c, hemoglobin A1c; WBC, white blood cell; Scr, serum creatinine; BUN, blood urea nitrogen; eGFR, estimated glomerular filtration rate; SOFA, Sequential Organ Failure Assessment; ACEI, angiotensin converting enzyme inhibitor; ARB, angiotensin II receptor blocker; RRT, renal replacement therapy.

### The association between postoperative GV and one-year mortality following CABG

Next, we examined whether postoperative glycemic variability (GV) may predict one-year mortality following CABG. Log-rank test showed that patients with higher postoperative GV had an increased one-year mortality rate compared to those with lower GV (log-rank p < 0.001, Figure 2).

**Figure 2.**
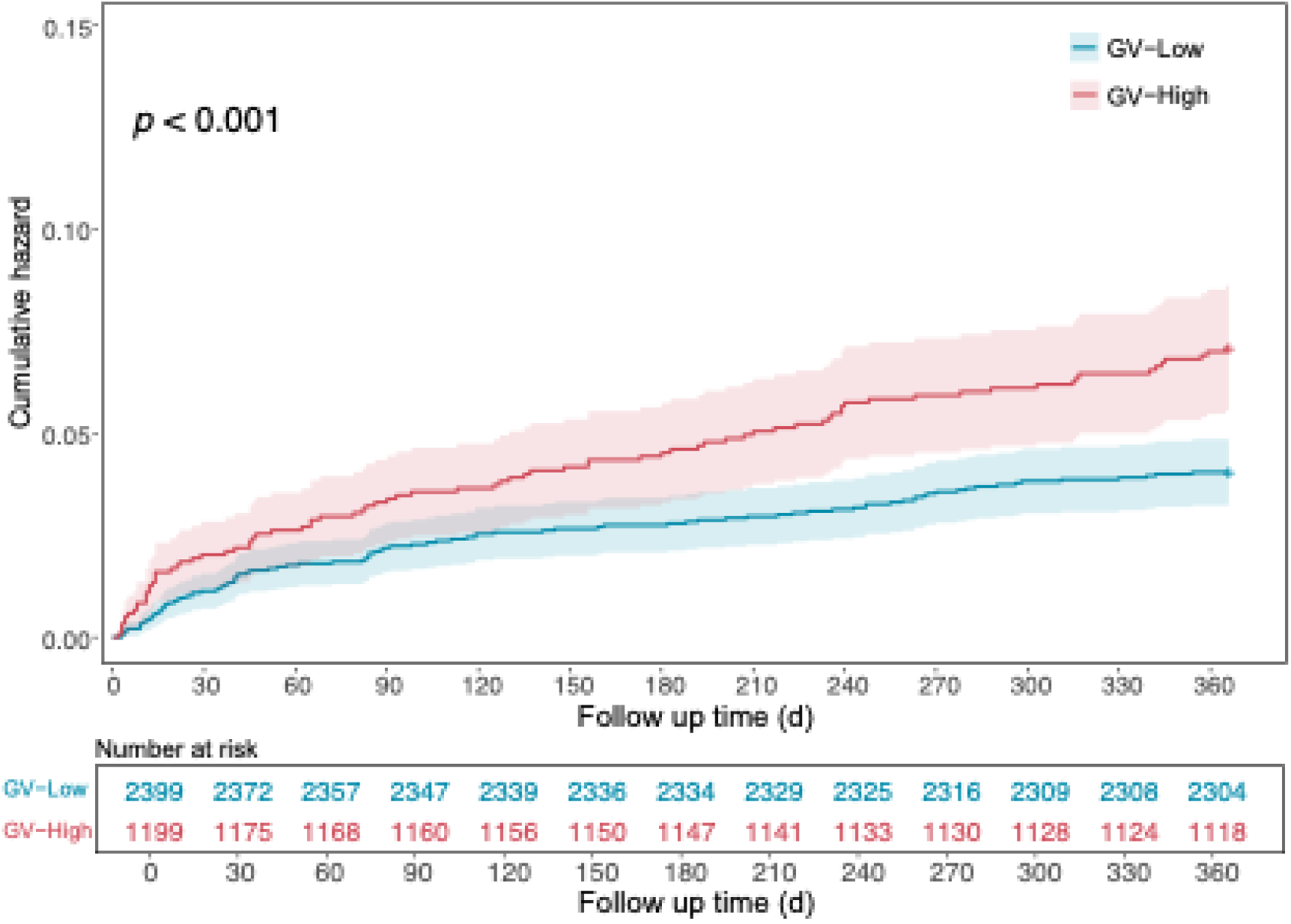
Kaplan–Meier analysis of one-year mortality following CABG. CABG, coronary artery bypass grafting; GV, glycemic variability. The GV-Low group was defined as GV < 0.219; The GV-High group was defined as GV ≥ 0.219.

Multivariate survival analysis with Cox’s regression model showed that postoperative GV was an independent risk factor for one-year mortality following CABG. An increase in postoperative GV was consistently associated with increased mortality risk across all models (Table 3). When GV was treated as a categorical variable, patients with high GV had significantly higher hazard ratio (HR) compared to those with low GV: HR of 1.751 (95% CI, 1.303–2.353) in the unadjusted model, HR of 1.665 (95% CI, 1.237– 2.242) in the partially adjusted model, and HR of 1.407 (95% CI, 1.039–1.906) in the fully adjusted model. When GV was analyzed as a continuous variable, each unit increase of GV was associated with significantly higher mortality risk, with HR of 10.372 (95% CI, 3.973–27.077) in the unadjusted model, 9.065 (95% CI, 3.387–24.263) in the partially adjusted model, and 4.599 (95% CI, 1.662–12.726) in the fully adjusted model. The *p* value for trend remained less than 0.05 across all models.

**Table 3.**
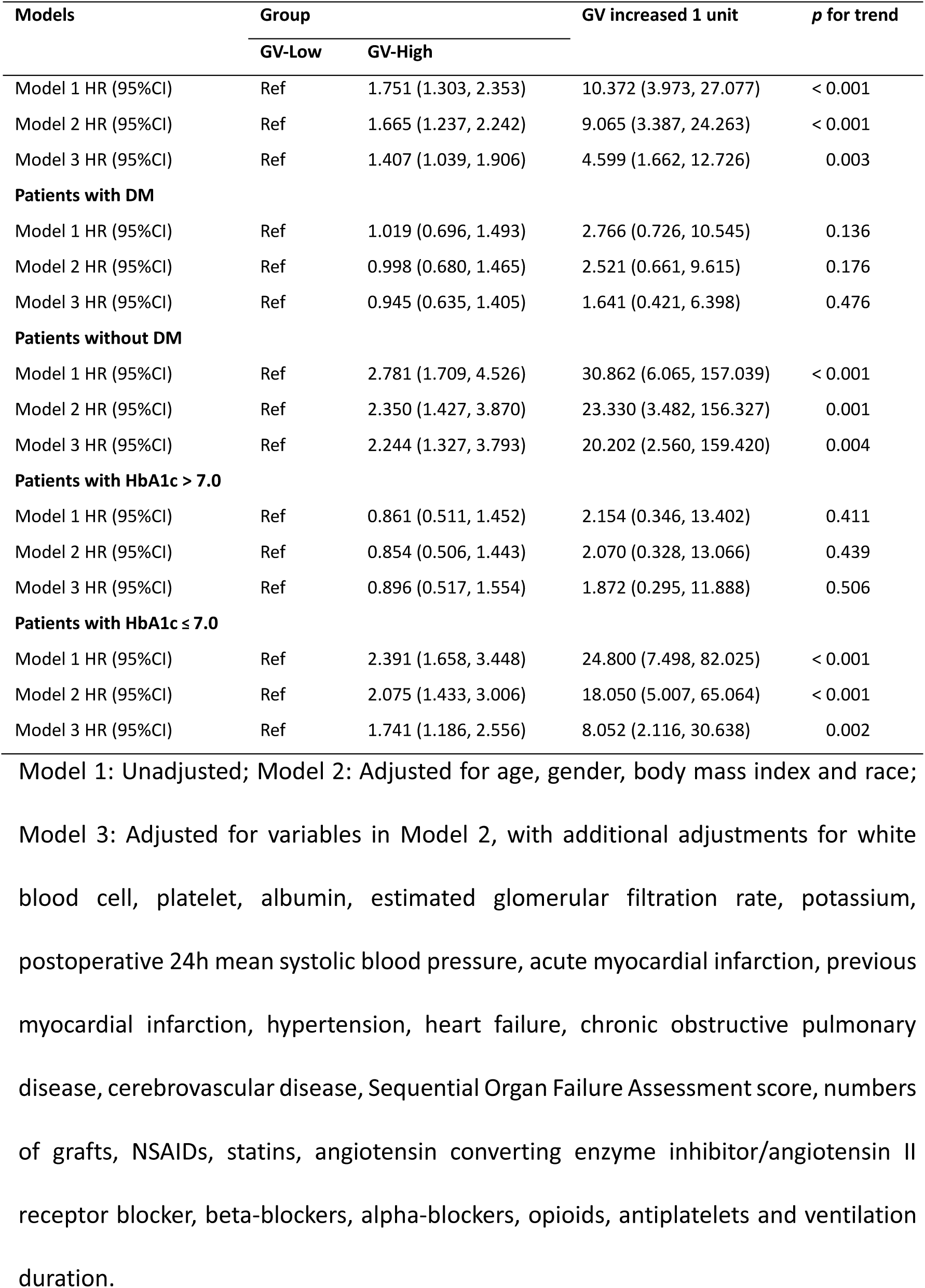
Cox proportional hazard ratio for one-year mortality following CABG.

| Models | Group |  | GV increased 1 unit | p for trend |
| --- | --- | --- | --- | --- |
|  | GV-Low | GV-High |  |  |
| Model 1 HR (95%CI) | Ref | 1.751 (1.303, 2.353) | 10.372 (3.973, 27.077) | < 0.001 |
| Model 2 HR (95%CI) | Ref | 1.665 (1.237, 2.242) | 9.065 (3.387, 24.263) | < 0.001 |
| Model 3 HR (95%CI) | Ref | 1.407 (1.039, 1.906) | 4.599 (1.662, 12.726) | 0.003 |
| **Patients with DM** |  |  |  |  |
| Model 1 HR (95%CI) | Ref | 1.019 (0.696, 1.493) | 2.766 (0.726, 10.545) | 0.136 |
| Model 2 HR (95%CI) | Ref | 0.998 (0.680, 1.465) | 2.521 (0.661, 9.615) | 0.176 |
| Model 3 HR (95%CI) | Ref | 0.945 (0.635, 1.405) | 1.641 (0.421, 6.398) | 0.476 |
| **Patients without DM** |  |  |  |  |
| Model 1 HR (95%CI) | Ref | 2.781 (1.709, 4.526) | 30.862 (6.065, 157.039) | < 0.001 |
| Model 2 HR (95%CI) | Ref | 2.350 (1.427, 3.870) | 23.330 (3.482, 156.327) | 0.001 |
| Model 3 HR (95%CI) | Ref | 2.244 (1.327, 3.793) | 20.202 (2.560, 159.420) | 0.004 |
| **Patients with HbA1c > 7.0** |  |  |  |  |
| Model 1 HR (95%CI) | Ref | 0.861 (0.511, 1.452) | 2.154 (0.346, 13.402) | 0.411 |
| Model 2 HR (95%CI) | Ref | 0.854 (0.506, 1.443) | 2.070 (0.328, 13.066) | 0.439 |
| Model 3 HR (95%CI) | Ref | 0.896 (0.517, 1.554) | 1.872 (0.295, 11.888) | 0.506 |
| **Patients with HbA1c ≤ 7.0** |  |  |  |  |
| Model 1 HR (95%CI) | Ref | 2.391 (1.658, 3.448) | 24.800 (7.498, 82.025) | < 0.001 |
| Model 2 HR (95%CI) | Ref | 2.075 (1.433, 3.006) | 18.050 (5.007, 65.064) | < 0.001 |
| Model 3 HR (95%CI) | Ref | 1.741 (1.186, 2.556) | 8.052 (2.116, 30.638) | 0.002 |

Model 1: Unadjusted; Model 2: Adjusted for age, gender, body mass index and race;
Model 3: Adjusted for variables in Model 2, with additional adjustments for white blood cell, platelet, albumin, estimated glomerular filtration rate, potassium, postoperative 24h mean systolic blood pressure, acute myocardial infarction, previous myocardial infarction, hypertension, heart failure, chronic obstructive pulmonary disease, cerebrovascular disease, Sequential Organ Failure Assessment score, numbers of grafts, NSAIDs, statins, angiotensin converting enzyme inhibitor/angiotensin II receptor blocker, beta-blockers, alpha-blockers, opioids, antiplatelets and ventilation duration.

In addition, a restricted cubic spline (RCS) regression model was used to investigate the relationship between the postoperative GV and the risk of one-year mortality following CABG. The RCS regression curve demonstrated a linear association between the GV and the one-year mortality risk after adjusting for confounding factors (*p*-non-linearity = 0.962, Figure 3). Further subgroup and interaction analyses were also conducted to investigate the relationship between the GV and the one-year mortality risk, revealing significant interactions with both DM status and preoperative HbA1c levels (Table 3 and Figure 4). Specifically, the association between the GV and the one-year mortality risk was stronger in patients without DM (odds ratio [OR], 2.781; 95% CI, 1.709–4.526). A similar strong association was observed in patients with preoperative HbA1c levels ≤ 7.0% (OR, 2.225; 95% CI, 1.537–3.221). In contrast, no significant association was found in patients with DM or those with preoperative HbA1c levels > 7.0% (Table 3 and Figure 4).

**Figure 3.**
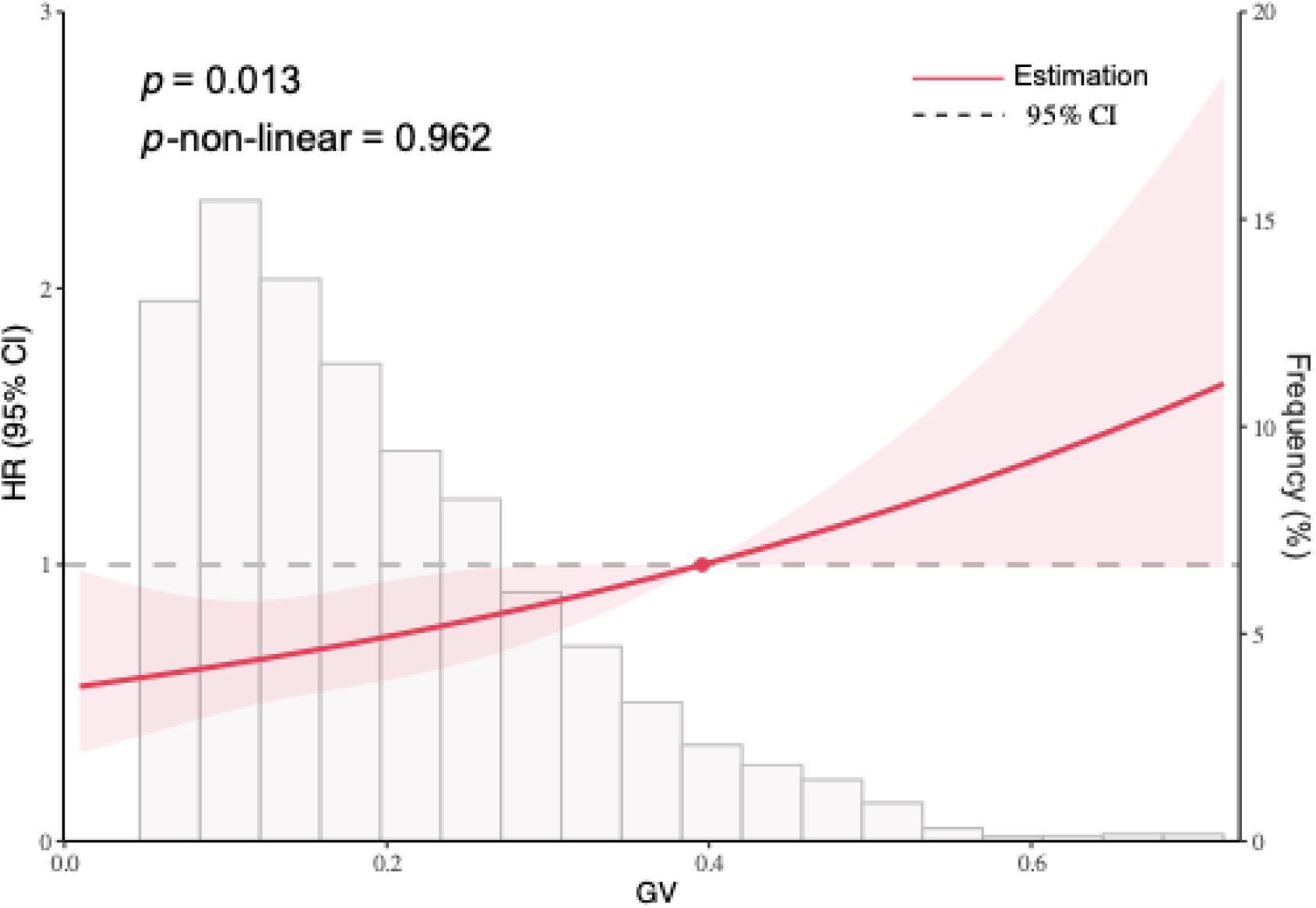
Restricted cubic spline analysis o of one-year mortality after CABG based on glycemic variability levels. The solid line represented the adjusted HR estimates, while the shaded area indicated the 95% CI. The horizontal dashed line denoted an HR of 1.0. GV, glycemic variability; CABG, coronary artery bypass grafting; HR, hazard ratio; CI, confidence interval.

**Figure 4.**
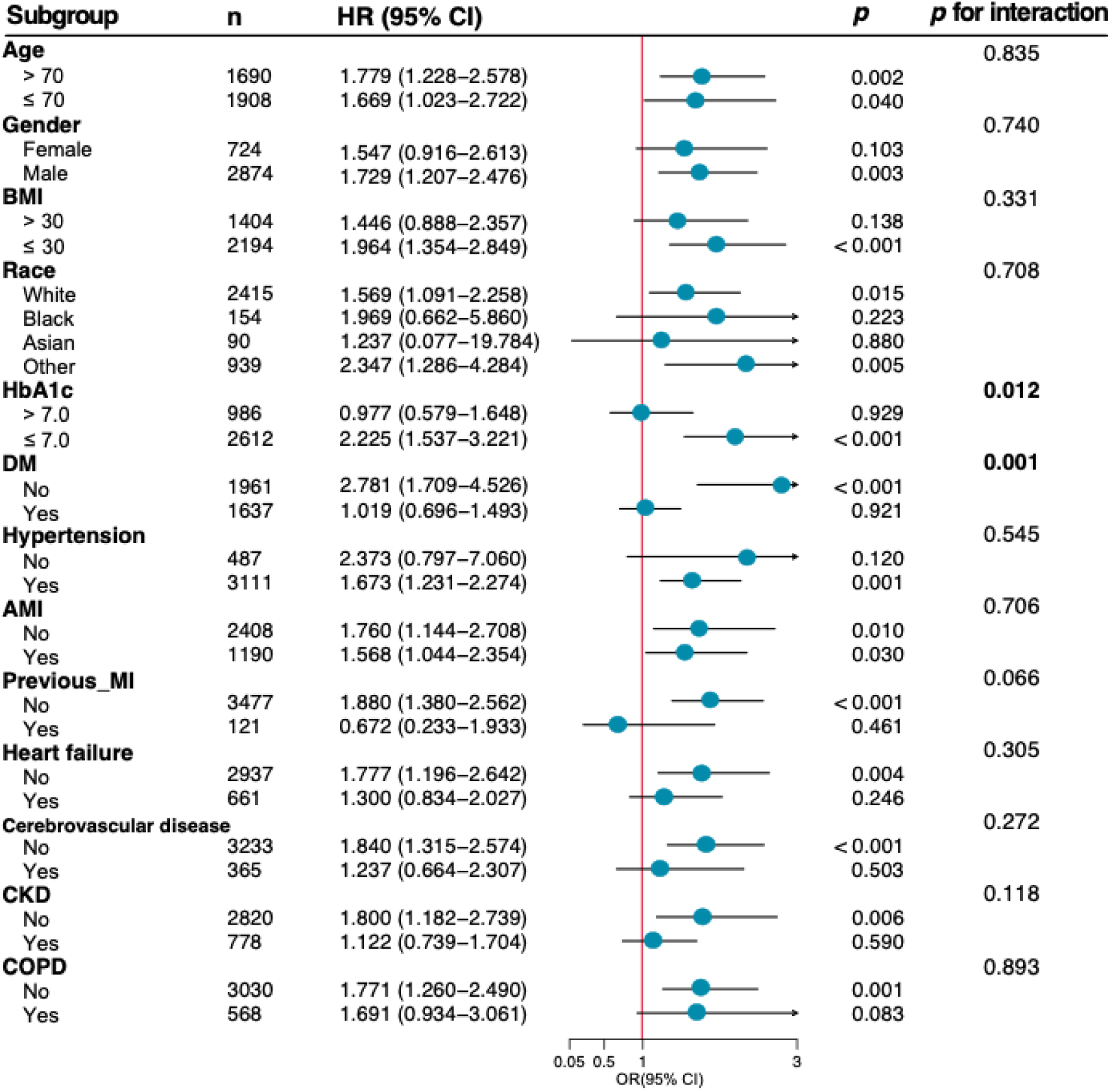
Forest plots illustrating the primary outcome among various subgroups. BMI, body mass index; HbA1c, hemoglobin A1c; DM, diabetes mellitus; AMI, acute myocardial infarction; MI, myocardial infarction; COPD, chronic obstructive pulmonary disease; CKD, chronic kidney disease; HR, hazard ratio; CI, confidence interval; OR, odds ratio.

### The association between postoperative GV and one-year mortality risk in patients with or without DM

In DM patients, the cumulative hazard curves indicated a slightly increased one-year mortality risk in the GV-High group compared to the GV-Low group (Figure 5A). However, RCS analysis revealed no significant association between GV and one-year mortality in this subgroup (*p* = 0.328; *p*-non-linear = 0.212) (Figure 6A). Similarly, Cox regression models did not identify a significant relationship, no matter whether GV was analyzed as a categorical variable (HRs, 0.945; 95% CI, 0.635–1.405) or as a continuous variable (HR per unit increase, 1.641; 95% CI, 0.421–6.398; *p* for trend = 0.476) (Table 3).

**Figure 5.**
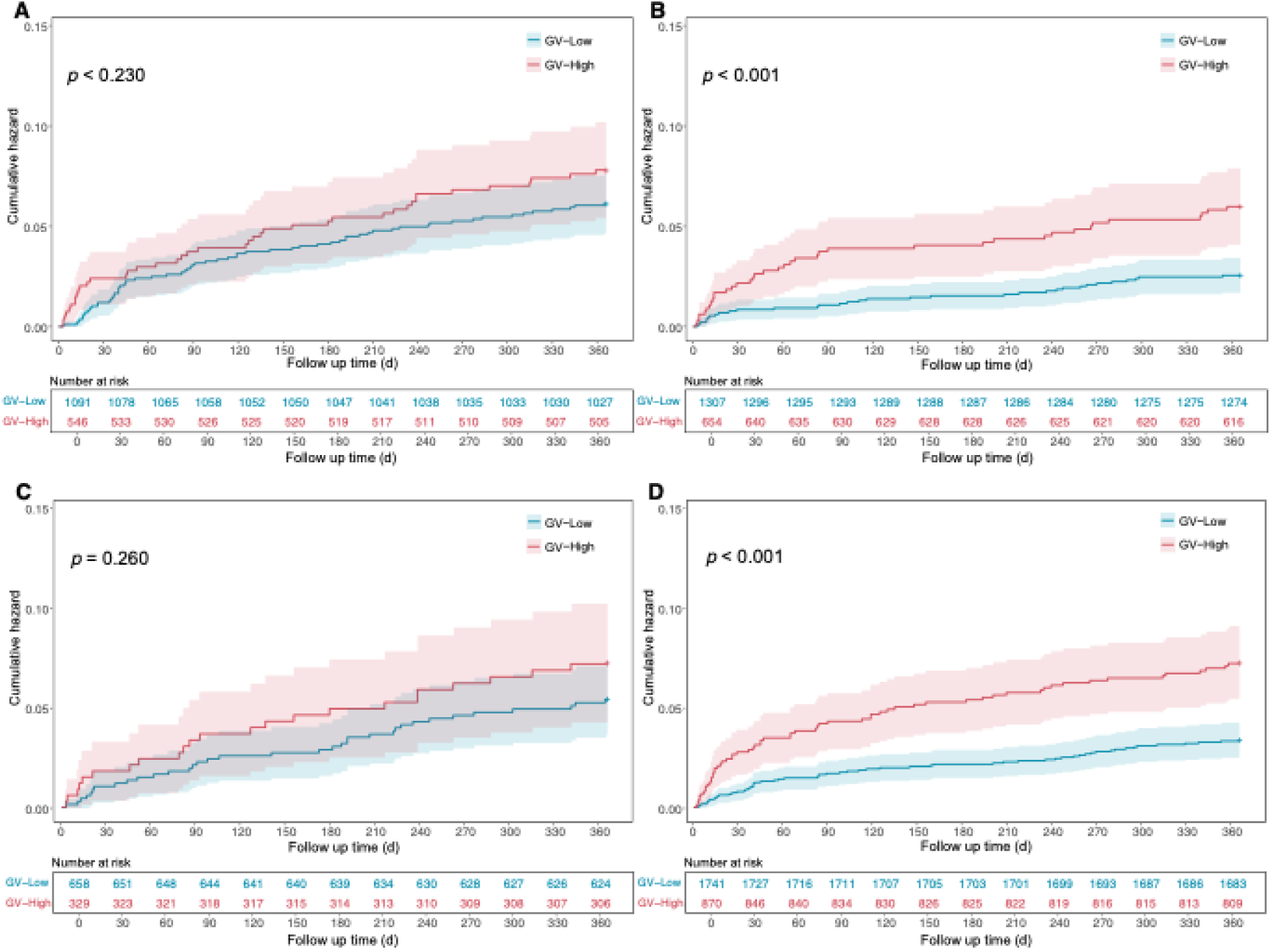
Kaplan–Meier analysis of one-year mortality following CABG based on glycemic variability levels in different subgroups. Kaplan-Meier curves illustrating the cumulative incidence of one-year mortality following CABG, based on high and low GV in patients with DM (**A**), in patients without DM (**B**), in patients with preoperative HbA1c > 7.0% (**C**), and in patients with preoperative HbA1c ≤ 7.0% (**D**). GV, glycemic variability; CABG, coronary artery bypass grafting; DM, diabetes mellitus; HbA1c, hemoglobin A1c.

**Figure 6.**
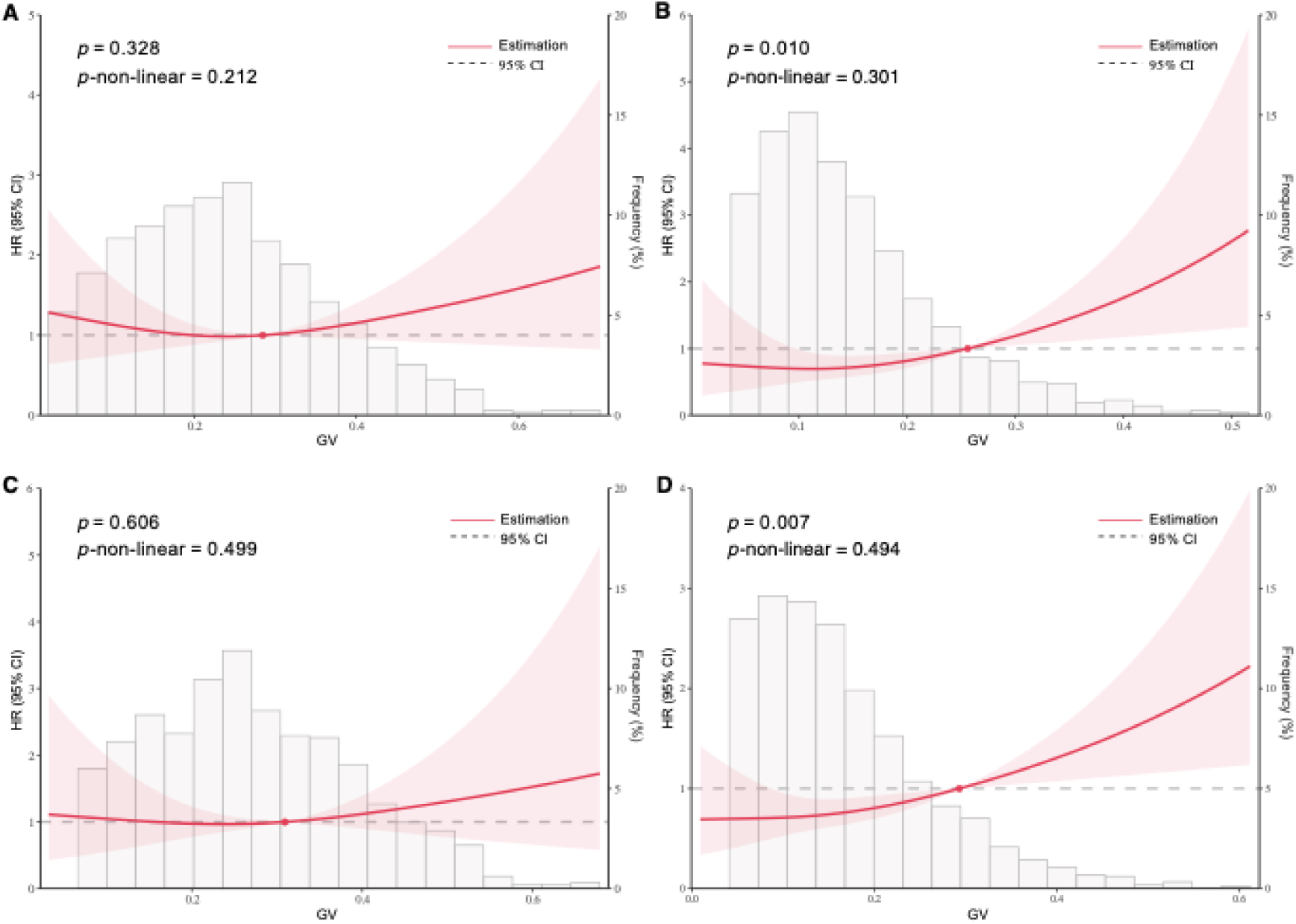
Restricted cubic spline analysis of one-year mortality following CABG based on glycemic variability in different subgroups. Subgroup analyses were conducted for patients with DM (**A**), patients without DM (**B**), patients with preoperative HbA1c > 7.0% (**C**), and patients with preoperative HbA1c ≤ 7.0% (**D**).The solid line represented the adjusted HR estimates, with the shaded area indicating the 95% CI. The horizontal dashed line denoted an HR of 1.0. GV, glycemic variability; CABG, coronary artery bypass grafting; DM, diabetes mellitus; HbA1c, hemoglobin A1c; HR, hazard ratio; CI, confidence interval.

In contrast, patients without DM exhibited a remarkedly higher one-year mortality risk in the GV-High group compared to the GV-Low group (Figure 5B). The RCS analysis showed a significant linear association (*p* = 0.010; *p*-non-linear = 0.301) (Figure 6B). The Cox regression analysis also supported this finding, demonstrating that an increased GV was significantly associated with an increased risk of one-year mortality (HR, 2.244; 95% CI, 1.327–3.793). Moreover, each unit increase in GV corresponded to an HR of 20.202 (95% CI, 2.560–159.420; *p* for trend = 0.004) (Table 3).

### The association between postoperative GV and one-year mortality risk in patients with different levels of perioperative HbA1c

In patients with HbA1c > 7.0%, Kaplan-Meier curves showed a slight increase in one-year mortality in the high-GV group compared to the low-GV group (Figure 5C). RCS analysis also showed no significant association (*p* = 0.606) or non-linear relationship (*p*-non-linear = 0.499) between GV and one-year mortality risk in patients with HbA1c > 7.0% (Figure 6C). Similarly, Cox regression models demonstrated no significant association regardless whether GV was treated as a categorical variable (HR, 0.896; 95% CI, 0.517–1.554) or as a continuous variable (HR per unit increase, 1.872; 95% CI, 0.295–11.888; *p* for trend = 0.506) (Table 3).

In contrast, in patients with HbA1c ≤ 7.0%, Kaplan-Meier curves (Figure 5D) and RCS analysis (Figure 6D) showed a strong positive association between the GV and the one-year mortality risk following CABG. The RCS model also demonstrated a significant relationship (*p* = 0.007), with no evidence of a non-linear association (*p*-non-linear = 0.494, Figure 6D). Cox regression models further supported this finding, showing that the increased GV was associated with the increased one-year mortality risk (HR, 1.741; 95% CI, 1.186–2.556), and each unit increase in GV corresponded to an HR of 8.052 (95% CI, 2.116–30.638; *p* for trend = 0.002) (Table 3).

### Sensitivity analysis of association between the postoperative GV and one-year mortality

To further assess the reliability of the postoperative GV in predicting the one-year mortality following CABG, a sensitivity analysis was performed (Table 4). The cohort of 3,598 patients was divided into two groups based on the GV median below or above with 1,799 patients in each group. Multivariable Cox regression analysis showed that patients with GV above the median had a higher one-year mortality risk than those with GV below the median (adjusted HR, 1.454; 95% CI, 1.059–1.997; *p* = 0.003). Subgroup analyses further revealed that this association was significant in patients without DM (adjusted HR, 1.755; 95% CI, 1.062–2.901; *p* = 0.008) or patients with HbA1c ≤ 7.0% (adjusted HR, 1.504; 95% CI, 1.026–2.204; *p* = 0.003), while no significant association was observed in patients with DM (*p* = 0.476) or those with HbA1c > 7.0% (*p* = 0.501).

**Table 4.** Association between postoperative glycemic variability and one-year mortality based on the glycemic variability median.

|  | GV below median | GV above median | Unadjusted HR (95% CI) | *p* | Adjusted* HR (95% CI) | *p* |
| --- | --- | --- | --- | --- | --- | --- |
| **Total population** | n=1799 | n=1799 |  |  |  |  |
| one-year mortality | 64 (3.557%) | 113 (6.281%) | 1.790 (1.317, 2.432) | < 0.001 | 1.454 (1.059, 1.997) | 0.003 |
| **With DM** | n=497 | n=1140 |  |  |  |  |
| one-year mortality | 31 (6.237%) | 75 (6.579%) | 1.052 (0.692, 1.599) | 0.136 | 0.924 (0.598, 1.425) | 0.476 |
| **Without DM** | n=1302 | n=659 |  |  |  |  |
| one-year mortality | 33 (2.535%) | 38 (5.766%) | 2.319 (1.455, 3.697) | < 0.001 | 1.755 (1.062, 2.901) | 0.008 |
| **HbA1c > 7.0%** | n=238 | n=749 |  |  |  |  |
| one-year mortality | 14 (5.882%) | 44 (5.874%) | 0.991 (0.543, 1.808) | 0.411 | 0.914 (0.493, 1.693) | 0.501 |
| **HbA1c ≤ 7.0%** | n=1561 | n=1050 |  |  |  |  |
| one-year mortality | 50 (3.203%) | 69 (6.571%) | 2.092 (1.454, 3.011) | < 0.001 | 1.504 (1.026, 2.204) | 0.003 |

\*Adjusted for age, gender, for age, gender, body mass index, and race, white blood cell, platelet, albumin, estimated glomerular filtration rate, potassium, postoperative 24h mean systolic blood pressure, acute myocardial infarction, previous myocardial infarction, hypertension, heart failure, chronic obstructive pulmonary disease, cerebrovascular disease, Sequential Organ Failure Assessment score, numbers of grafts, NSAIDs, statins, angiotensin converting enzyme inhibitor/angiotensin II receptor blocker, beta-blockers, alpha-blockers, opioids, antiplatelets, ventilation duration.

### Assessment of goodness-of-fit in comparison of models

The goodness-of-fit measure in models revealed that adding the postoperative GV to the baseline model significantly improved its performance, as indicated by reductions in both the Akaike information criterion (AIC) and Bayesian information criterion (BIC) values (Table 5). In non-DM patients or patients with HbA1c ≤ 7.0%, the inclusion of GV further reduced the AIC and BIC values and yielded significant likelihood ratio test (LRT) results (*p* = 0.005 and *p* = 0.008, respectively). These findings suggested that the postoperative GV is also a stronger predictor for one-year mortality following CABG.

**Table 5.** Assessment of the goodness-of-fit of model.

| | Patients without DM | | Patients with HbA1c $\leq 7.0$ | |
| --- | --- | --- | --- | --- |
|  | Model 3 | Model 3 + GV | Model 3 | Model 3 + GV |
| AIC | 994.410 | 988.594 | 1727.022 | 1721.881 |
| BIC | 1060.027 | 1056.475 | 1807.617 | 1805.255 |
| LRT | Reference | 7.815 | Reference | 7.141 |
| df | Reference | 1 | Reference | 1 |
| *p*-value | Reference | 0.005 | Reference | 0.008 |

DM, diabetes mellitus; GV, glycemic variability; AIC, Akaike information criterion; BIC,
Bayesian information criterion; LRT, Likelihood ratio test.

## Discussion

This study found postoperative GV as an independent predictor of one-year mortality following coronary artery bypass grafting (CABG), with increased postoperative GV associated with higher mortality risk. Notably, the one-year mortality risk was also increased in patients without diabetes mellitus (DM) or patients with preoperative glycated hemoglobin (HbA1c) levels ≤ 7.0%. These findings suggest the importance of strategies to reduce postoperative GV, particularly in non-DM patients and those with lower HbA1c, to enhance survival outcomes after cardiac surgery.

Chronic heart disease (CAD), a multifactorial and life-threating condition, remains the leading cause of death in the United States or worldwide ^14^. The CABG, which restores blood flow and oxygen to the heart by addressing blockages or narrowing in the coronary arteries, is the preferred treatment for CAD ^1^. However, large-scale studies in the United States reported an in-hospital mortality rate of 3.4% among 3,374,743 CABG patients ^3^. In the current study, the follow-up period was extended to postoperative one year, showing a slightly higher overall mortality rate of 4.92%, comparable to the findings from another cohort study (2008–2016) reporting a one-year postoperative mortality rate of 4.62% ^15^. Given the high postoperative mortality rate, it is crucial to identify high-risk factors and implement preventive strategies to reduce mortality following CABG.

Stress is a key contributor to postoperative insulin resistance and hyperglycemia ^16^, which are associated with increased short-term mortality following cardiac surgery ^9,17–20^. However, assessing perioperative glycemic changes is challenging due to the lack of recorded preoperative baseline glycemic levels. Some studies suggested that postoperative GV may serve as a reliable metric for capturing dynamic changes in perioperative glycemic profiles ^21–23^. A single-center observational cohort study previously reported that increased GV in patients with elevated preoperative HbA1c levels was associated with higher rate of 30-day postoperative adverse outcomes following CABG, including myocardial infarction (MI), reoperations, sternal infections, cardiac tamponade, pneumonia, stroke, or renal failure, but not with in-hospital mortality ^9^. In contrast, our study found that increased postoperative GV was significantly associated with higher one-year mortality following CABG. This was further verified by two adjusted models of Cox’s regression. A possible explanation for this discrepancy is that our study included a larger cohort of 3,598 patients and extended the follow-up period to 1 year, providing a more comprehensive evaluation of long-term outcomes.

The underlying mechanisms by which GV influences postoperative mortality remain unclear, but several hypotheses may be proposed. One key theory suggests that fluctuations in glucose levels contribute to oxidative stress by promoting the excessive production of free radicals, overwhelming the cells’ intrinsic antioxidant defense systems. This oxidative imbalance may damage cellular structures and impair physiological function ^21^. Additionally, GV has been implicated in triggering apoptosis in human umbilical vein endothelial cells, contributing to endothelial dysfunction, a critical factor in vascular health ^24^. Furthermore, GV may play a role in the development of neuropathy with striking axonal atrophy of chiefly neuronal fibers ^24^.

These findings underscore the potential systemic impact of GV, highlighting the need for targeted strategies to mitigate its effects, particularly in the context of cardiac surgery.

We further conducted subgroup and interaction analysis to investigate the relationship between postoperative GV and one-year mortality risk, and found that the diabetes status may play a crucial role in this association. Surprisingly, in non-DM patients, increased GV was significantly associated with a higher risk of one-year mortality, whereas no statistically significant association was observed in DM patients. There might be a key signal indicating that the DM patients may develop long-term physiological adaptations (conditioning effects) to cope with blood glucose fluctuations, while the non-DM patients are more vulnerable to the metabolic burden of glycemic swings, thereby increasing their mortality risk ^25–27^. Moreover, stringent glycemic control (80–110 mg/dL) has been shown to lower infection risk and improve short-term outcomes following surgery, but may also increase the risk of hypoglycemia, a condition particularly harmful in DM patients ^26,28^. Conversely, more liberal glycemic targets (140–180 mg/dL) reduced hypoglycemic risk but may increase GV, potentially leading to adverse outcomes in non-DM individuals ^29,30^.

Furthermore, we found that increased GV after CABG, particularly in patients with preoperative HbA1c ≤ 7.0%, had a high rate of one-year mortality, regardless of diabetic status. This work expands on a previous finding that demonstrated the relationship between preoperative blood glucose control and major adverse events (MAEs), suggesting the importance of postoperative GV after cardiac surgery. Balachundhar et al. reported that postoperative GV was increased in patients with poor preoperative glycemic control (HbA1C ≥ 6.5%), thereby associated with MAEs following CABG ^9^. However, Kathleen et al. found that an increased 24-h but not 12-h postoperative GV after CABG was a predictor of MAEs, and preoperative HbA1c was not associated with MAEs after adjusting for postoperative mean glucose and GV ^31^. Similarly, a large retrospective cohort including 431,480 patients demonstrated that though HbA1C was positively associated with perioperative glucose, but not associated with increased 30-day mortality after controlling for glucose ^32^. These discrepancies may stem from the inherent limitations of HbA1c, which only reflects mean glycaemia over the preceding 8–12 weeks and fails to capture acute glycaemia fluctuations^33^. Indeed, even patients with the same HbA1c levels may experience markedly different complication rate^21,34,35^. Importantly, acute glucose fluctuations can induce oxidative stress, inflammation, endothelial dysfunction, and altered gene expression, leading to adverse outcomes^22,35,36^. These findings highlight the need for individualized glycemic management strategies based on pre-diabetes or diabetes status to optimize surgical outcomes.

This study has several limitations. Firstly, due to the absence of continuous glycemic monitoring data in the MIMIC database, we used blood glucose records from the first 72-h postoperative period to calculate GV and this approach might introduce a certain degree of bias. Secondly, as a retrospective study, our analysis was constrained by the data available in the MIMIC database. Despite performing multivariable adjustments and subgroup analyses, residual confounding factors might impact clinical outcomes. Notably, the potential confounders, such as admission status and the use of cardiopulmonary bypass, were unavailable in the database. Finally, the patients included in this study were from a single center in the United States, which might limit the generalizability of the findings. These observational conclusions should be verified through further high-quality randomized controlled trials.

## Conclusion

In conclusion, postoperative glycemic variability (GV) after coronary artery bypass grafting is an independent predictor of one-year mortality in patients without diabetes mullites or with preoperative HbA1c ≤ 7.0%. Our findings highlight increased postoperative GV is associated with a substantially increased risk of postoperative mortality, emphasizing the need for careful glycemic management in this patient population.

## Data Availability

Publicly available datasets were analyzed in this study. These data can be found at https://mimic.mit.edu/.

## Acknowledgements

The authors have declared that no competing interest exists.

